# Geospatially-resolved public-health surveillance via wastewater sequencing

**DOI:** 10.1101/2023.05.31.23290781

**Authors:** Braden T Tierney, Jonathan Foox, Krista A Ryon, Daniel Butler, Namita Damle, Benjamin G Young, Christopher Mozsary, Kristina M. Babler, Xue Yin, Yamina Carattini, David Andrews, Natasha Schaefer Solle, Naresh Kumar, Bhavarth Shukla, Dusica Vidovic, Benjamin Currall, Sion L. Williams, Stephan C. Schürer, Mario Stevenson, Ayaaz Amirali, Cynthia C. Beaver, Erin Kobetz, Melinda M. Boone, Brian Reding, Jennifer Laine, Samuel Comerford, Walter E. Lamar, John J. Tallon, Jeremy Wain Hirschberg, Jacqueline Proszynski, Mark E. Sharkey, George M Church, George S Grills, Helena M. Solo-Gabriele, Christopher E Mason

## Abstract

Wastewater, which contains everything from pathogens to pollutants, is a geospatially-and temporally-linked microbial fingerprint of a given population. As a result, it can be leveraged for monitoring multiple dimensions of public health across locales and time. Here, we integrate targeted and bulk RNA sequencing (n=1,419 samples) to track the viral, bacterial, and functional content over geospatially distinct areas within Miami Dade County from 2020-2022. First, we used targeted amplicon sequencing (n=966) to track diverse SARS-CoV-2 variants across space and time, and we found a tight correspondence with clinical caseloads from University students (N = 1,503) and Miami-Dade County hospital patients (N = 3,939 patients), as well as an 8-day earlier detection of the Delta variant in wastewater vs. in patients. Additionally, in 453 metatranscriptomic samples, we demonstrate that different wastewater sampling locations have clinically and public-health-relevant microbiota that vary as a function of the size of the human population they represent. Through assembly, alignment-based, and phylogenetic approaches, we also detect multiple clinically important viruses (e.g., *norovirus*) and describe geospatial and temporal variation in microbial functional genes that indicate the presence of pollutants. Moreover, we found distinct profiles of antimicrobial resistance (AMR) genes and virulence factors across campus buildings, dorms, and hospitals, with hospital wastewater containing a significant increase in AMR abundance. Overall, this effort lays the groundwork for systematic characterization of wastewater to improve public health decision making and a broad platform to detect emerging pathogens.

## Introduction

Wastewater is a wellspring for geospatially-delineated, epidemiologically-relevant public-health data. One sample contains a cross-sectional collection of human pathogens, commensals, animal/plant detritus, and environmental features (e.g., pollutants) deriving from a specific locale^1^. Depending on sampling location, it represents small populations, like the residents of one building or entire cities^2^. As a result, wastewater is a potential source for continuous and precise monitoring of public health for communities of different sizes^3^. Applications of wastewater-based epidemiological surveillance include tracking community drug abuse, pollutants, and pathogen load^4–6^.

The SARS-CoV-2 pandemic galvanized efforts to develop low-cost tools for pathogen surveillance. Targeted sequencing of SARS-CoV-2 wastewater abundance gained popularity for its advantages over clinical or individual testing-based approaches^7^. Wastewater surveillance 1) can predict outbreaks before cases spike^8^, 2) is not readily identifiable at the patient level, reducing privacy concerns, 3) captures undersampled populations that may not be regularly tested^9^, 4) is more cost-effective than doing repeated individual tests^10^, and 5) reduces the reliance on self-reported data, which has proven a challenge in estimating both global SARS-CoV-2 burden and individual vaccination status^11^.

However, the public health utility of wastewater surveillance extends beyond monitoring single pathogens. Many past wastewater studies tracked environmental pollutants, like heavy metals and fertilizer runoff^12^. Recent efforts have used a combination of targeted (e.g. Polymerase-Chain-Reaction(PCR)-based) and untargeted (e.g. shotgun) amplification approaches to characterize multiple pathogen abundances simultaneously, including clinically-relevant enteric viruses^13^. These have revealed uncharacterized microbial life in wastewater, some of which has the potential to be biomarkers for aspects of human and societal health. In total, combining targeted pathogen tracking with broad monitoring of epidemiologically relevant microbial and environmental signals, pandemics and other public health crises can be better monitored and potentially mitigated.

While the potential impact of wastewater surveillance is clear, there are key gaps that must be addressed before its full public health potential can be realized. These include (1) standardization and development of sampling protocols, (2) analytic workflow construction, and (3) biomarker discovery and validation. The development of consistent protocols for sampling, sequencing, and analyzing wastewater data is critical for it to achieve any modicum of clinical impact. Further, once data standards are established, the global ecology of geospatially varying, longitudinal wastewater samples must be characterized and, in turn, associated with high-priority public health relevant features, from pollutant levels to disease. Without a basic understanding of the pan-domain microbial content of consistently processed wastewater, the ability to make policy decisions based on its composition will be hindered.

As a step towards public health-relevant wastewater surveillance, as well as to demonstrate the utility of uniting targeted and untargeted sequencing, we collected and analyzed 966 (weekly, on average) samples from 26 sites in Miami-Dade County between 2020 and 2022 (Fig 1A, Table 1). We performed both targeted sequencing to estimate SARS-CoV-2 burden and, on a subset of samples (N = 453), we completed bulk RNA sequencing to ascertain the broader microbial community. We sampled diverse locations, some representing small populations (dormitories and hospitals with a few to many hundreds of residents plus a regional wastewater treatment plant representing a population of over 800,000). We developed standardized collection and analysis workflows to holistically evaluate the microbial content of wastewater and generate public-health relevant guidance (Fig 1B)^7, 14–17^.

**Figure 1:**
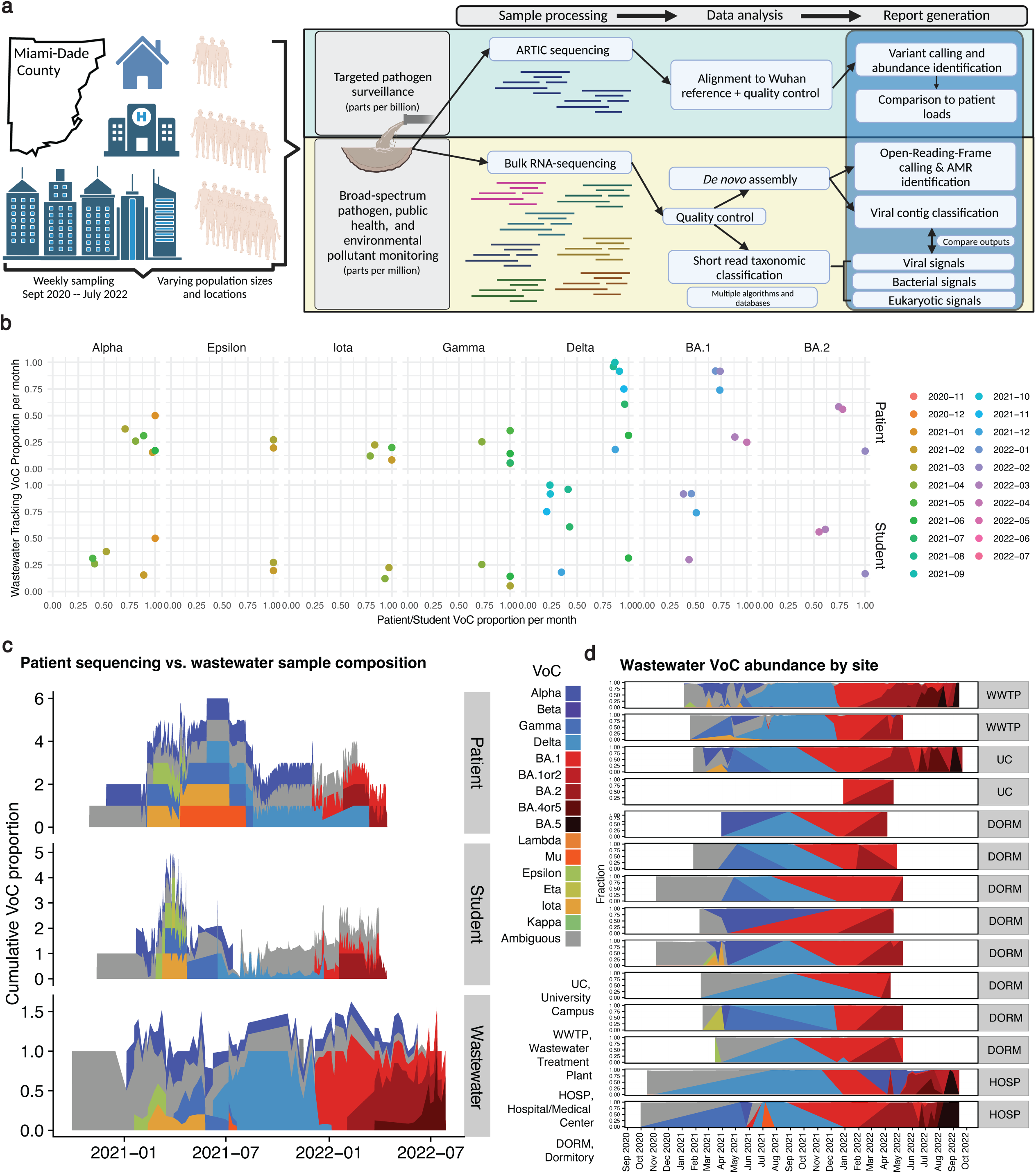
Overview of approach and targeted sequencing of SARS-CoV-2. A) Samples were taken at (on average) weekly intervals between 2020 and 2022 from 26 sites within Miami-Dade County. Samples were sequenced with targeted ARTIC sequencing to measure SARS-CoV-2 abundance and bulk RNA-sequencing to ascertain the broader microbial community. A variety of algorithms and analytic approaches were employed to identify and compare the taxonomic and functional profiles of each site across time and space, with the end result being a systematically characterized dataset with comparisons to clinical data to provide information relevant to public health surveillance showing the evolution of variants over space and time. B) The monthly VoC proportions across datasets. Point color corresponds to month. X-axis is the proportion of samples annotated as a given VoC for patient or student samples (derived from individual tests). Y-axis is the average variant abundance in targeted wastewater sequencing. C) An additional, density-plot-based, view of all variants in wastewater vs patient/student cohorts over time. D) The variation in wastewater VoCs across time in different sampling sites.

Bioinformatically, the analysis process involved quality control (i.e., filtering targeted sequencing data for samples with >75% SARS-CoV-2 genome coverage) as well as, in the case of bulk data, processing with multiple algorithmic workflows (i.e., k-mer based taxonomic classification, alignment-based taxonomy, *de novo* assembly-based approaches) and databases to yield multiple taxonomic and functional calls for each sample. The end result is a report integrating multiple views into the composition of each wastewater sample during various waves of the COVID-19 pandemic.

## Results

### Geospatially-resolved wastewater vs. clinical SARS-CoV-2 variant tracking

Like other efforts^8, 18^, we used targeted sequencing to generate high-resolution maps of SARS-CoV-2 Variant of Concern (VoC) levels across time based on coverage in wastewater sequencing data (Supp Fig 1, Supp Table 1). We observed across all wastewater samples the rise and fall of the VoCs that dominated the pandemic up to Omicron BA5. Wastewater VoC abundance temporally aligned with 5,442 sequenced or otherwise characterized samples from University of Miami (UM) students (N = 1,503) and the UM UHealth hospital patients (N = 3,939 patients) (Fig 1B-C).

The initial observation of different VoCs in wastewater was coincident with their initial identification in these other samples, especially data later in the pandemic as dominant variants became more defined. We additionally observed that the monthly proportion of VoCs in wastewater varied over time (Fig 1B-C), especially compared to the proportion of VoCs detected in the clinical and student settings (Fig 1B); in other words, wastewater VoCs appeared to track the monthly waves the pandemic more effectively that individual testing. Variant detection in wastewater preceded either (or both) student/patient detection for 5/8 VoCs identified in 2021 and 2022 (Supp Table 1). On average, VoCs in wastewater were identified within 1.8 days of their identification in either student or patient samples. Notably, the delta variant was detected eight days in wastewater prior to it appearing in student samples.

Wastewater data provided a more complete picture of all variants circulating in Miami-Dade County than either the student or patient data alone, as these approaches relied on random sampling based on which single individuals were tested (Fig 1C). For example, the lambda variant was identified in the wastewater and patient data (at high coverage for the clade-specific variants), but not in the student data, further indicating the propensity for individual testing to overlook potential circulating VoCs.

The continual sampling of different wastewater sites, however, enabled us to overcome the random bias generated by individual testing to observe variation in rarer variants across time (Fig 1D). For example, the Eta variant was present in March/April 2021 in the UM Campus Basin, whereas Lambda was more detectable in wastewater treatment plants and UM dormitory sites. Mu was observed in hospital wastewater but nowhere else. In this sense, geospatially resolved, continual targeted monitoring was found to provide transmission information for alternative variants from potentially survey-undersampled, non-hospital-associated communities for alternative variant transmission.

We found that wastewater based VoC surveillance enabled tracking the mutational-level transitions between dominant strains across time (Fig 2). We were able to observe transitions in both unique and recurrent mutations between VoCs. This high-resolution phylogenetic tracking enabled monitoring of the rise and fall of competing variants across space. We attained clear tracking of the variant mutational landscape despite using only weekly sampling.

**Figure 2:**
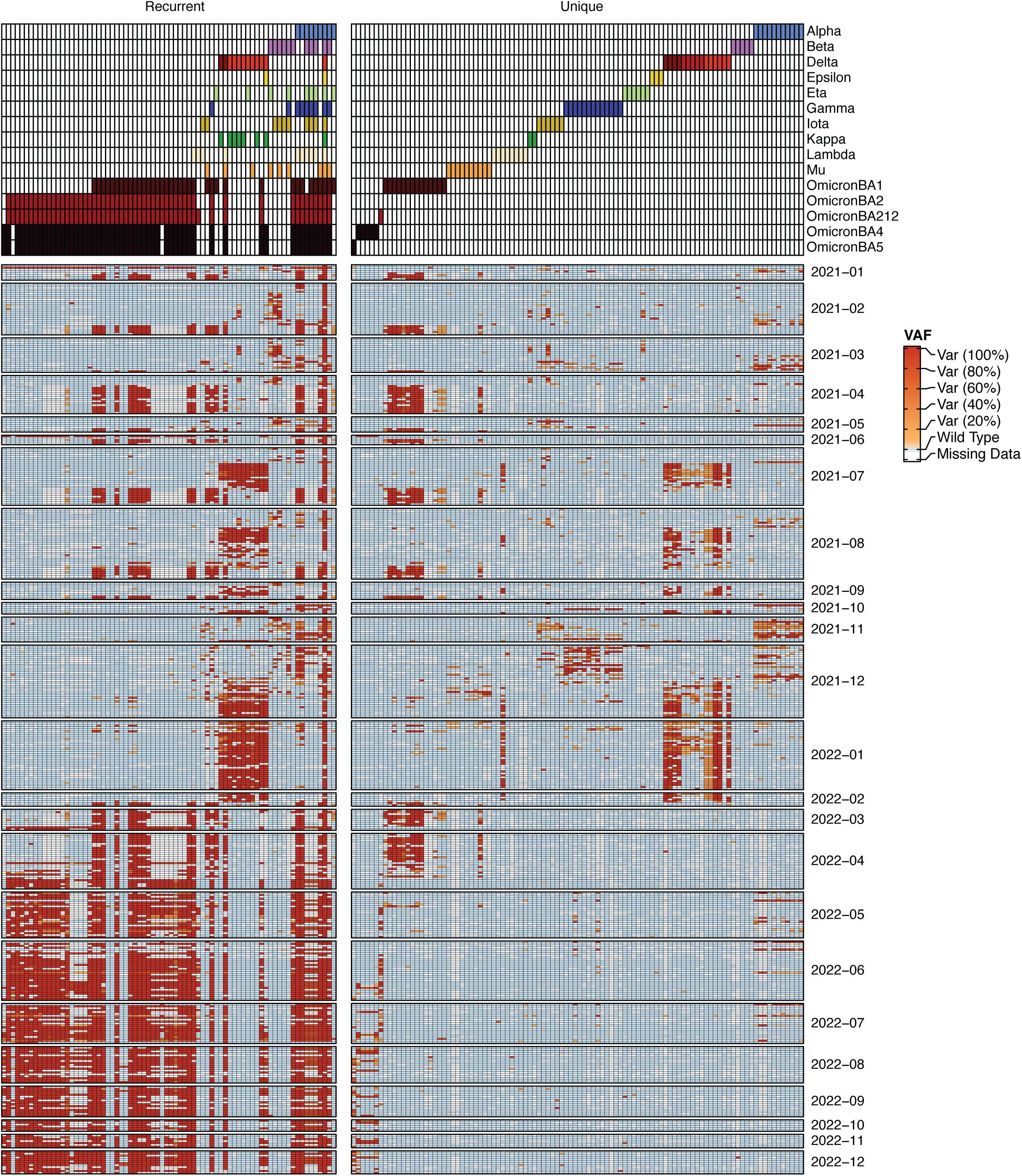
Tracking Variants of Concern (VoCs) across time. Right heatmap is unique mutations between different VoCs, left comprises recurrent mutations. We show samples starting when non-ambiguous variants were present in the wastewater data (post 2021).

### Public health tracking via wastewater-based monitoring of both pathogen and gut commensal activity

By comparison, bulk RNA sequencing captured a diverse array of pathogens as well as bioindicators for gut health. We compared multiple taxonomic annotation approaches and found complementary results in the profiles they produced (Supp Fig 2, Supp Table 2). A bacterial phylogeny demonstrated the high microbial diversity of wastewater (Fig 3A). Detected families included many human gut commensals (e.g. *Ruminococcaceae, Lachnospiraceae, Akkermansiaceae, Bifidobacteriaceae*) as well as oral microbiome commensals (e.g., *Nanogingivalaceae*, *Fusobacteriaceae*). Outside of host-associated organisms, many different environmental families (e.g. *Aquaspirilaceae, Rhodocyclaceae*) were also present. The amalgam of human and environmental microbes increased our confidence that it could potentially contain a broad spectrum of biomarkers indicative of different public health-relevant indications.

**Figure 3:**
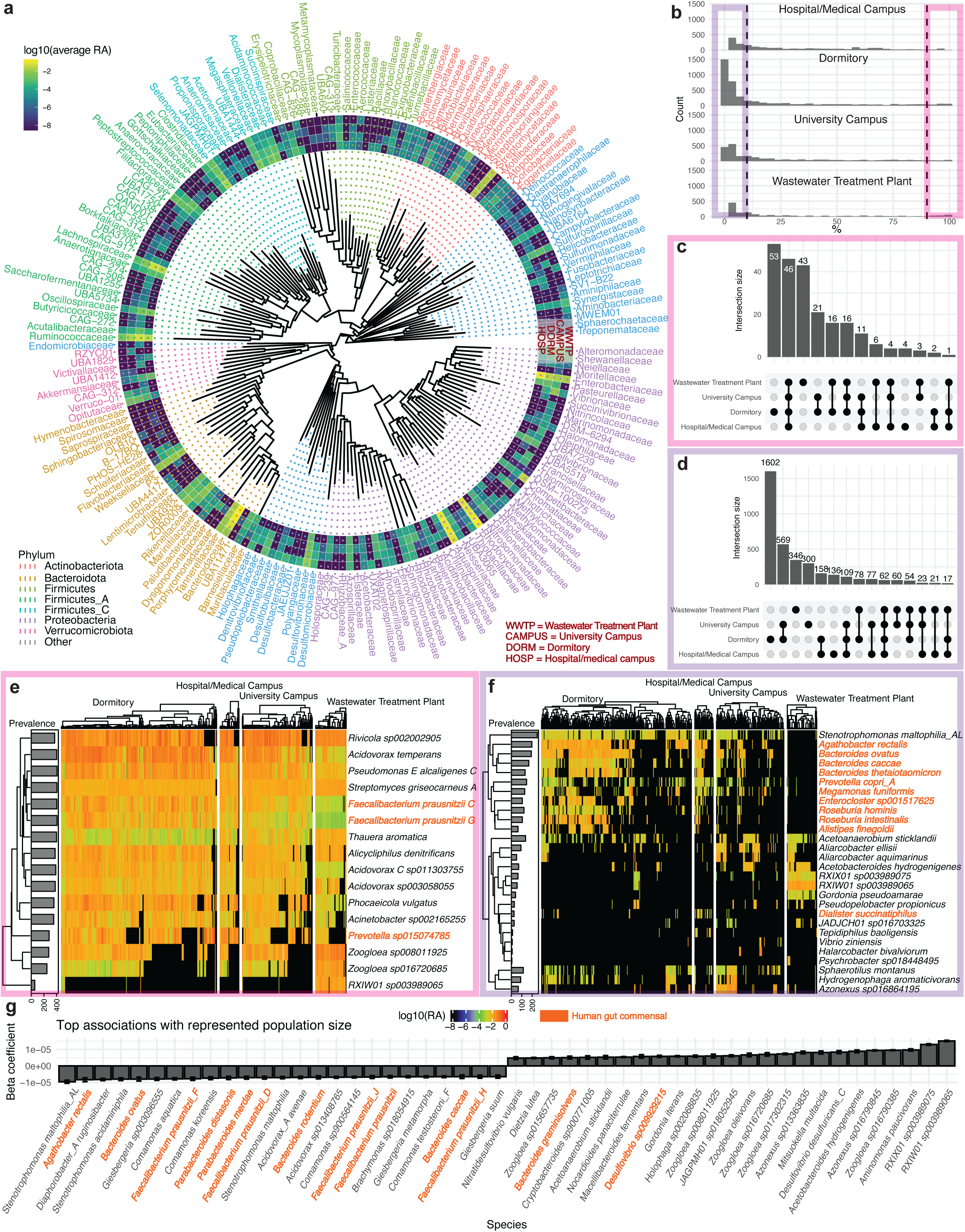
Wastewater bacterial phylogenetics and diversity. A) The average abundance of all bacterial families represented in the metatranscriptomic data. Outer rings correspond to mean abundance within a family across the different sampling site types. B) The prevalence of all identified bacterial species across different types of sampling locations. C-D) The overlap between high prevalence (C) and low prevalence (D) organisms across different sampling locations. E-F) The log10(relative abundance) of high-prevalence (E) and low-prevalence (F) bacterial species across all sites. These two panels display, at most, the union of the top 25 most abundant organisms across each sampling location and category (high or low prevalence). G) The top 25 most-and least-associated bacteria with represented population size of a given sampling location. For panels E-G), orange taxa indicate a gut commensal species. RA is an abbreviation of relative abundance.

The high diversity of wastewater yielded a large number of detected species and limited conservation across geographic locations. Microbial prevalence across all sites followed a left-skewed distribution (Fig 3B), with the few organisms being high prevalence (present in >90% of samples for a given sampling location) and the majority being low prevalence (present in <10% of samples). We identified “core” bacterial taxonomy within wastewater, comprising 46 species that were present in 90% of samples across all four types of sites (Fig 3C, Supp Table 2); these were predominantly environmental taxa. The 62 low prevalence taxa that overlapped between all sites were mainly host-associated species. Dormitories had the most diverse microbiomes, with 1,602 taxa identified uniquely in them, which may reflect the younger population’s microbiome.

High-prevalence bacteria (Fig 3E) were predominantly environmental organisms that had been documented as common in wastewater previously (e.g., *Acidovorax* species*, Rivicola* species, *Acinetobacter* species). However, some of the other species found in high prevalence and abundance were health-associated human gut commensals. These included the beneficial bacteria *Faecalibacterium praustinizii*, *Bacteroides uniformis, and Parabacteroides distasonis*^19^*. Alicycliphilus denitrificans* was also present across all sites; *A. denitrificans* is capable of reducing nitrate, which is a common groundwater pollutant monitored closely in wastewater.

Many low-prevalence bacteria were also host-associated (Fig 3F). These included *Bacteroides ovatus, Bacteroides caccae, Bacteroides thetaiotaomicron, Roseburia intestinalis, Enterocloster* species, *Prevotella copri*, *Alistipes finegoldii*, and *Dialister succinatiphilus*^19^. While pathogen detection dominates the wastewater field, the identification of beneficial commensals in high abundance indicates the inverse may also be true: precisely tracking the health and diet of a specific population. Notably, gut commensals were predominantly found in the dormitories as opposed to the wastewater treatment plants, further indicating the sensitivity of low-population sites to human-associated organisms.

Having observed the variation in diversity and content of microbiomes as a function of site, we hypothesized the population represented by a given locale would drive taxonomic content. To this end, we completed a Microbial Association Study (MAS) between species abundance and sampling sites, representative population size, and various environmental features. We found host-associated organisms were, indeed, statistically associated with smaller populations (Fig 3G, Supp Table 3). These associations highlighted opportunistic pathogens detectable in wastewater as well (e.g., *Stenotrophomonas maltophilia*). We additionally observed other statistically significant, environmental indicators of wastewater microbiome content (Supp Fig 3), including pH (71 positive associations, 61 negative), specific conductivity (8 positive, 129 negative), and dissolved oxygen content (184 positive associations, 68 negative). Notably, specific conductivity had positive associations with predominantly halophilic organisms, like *Cobetia marina* and *Halomonas E sp014840935*. In contrast to our prior work, there were no association found for viral load^20^.

We additionally performed a more targeted search for additional potential bacterial pathobionts of interest. We used the most sensitive taxonomic classifier (kraken2) for this, as it was most likely to pick up low abundance organisms; however, it is also prone to false positives, so any results would need to be validated with further effort. Regardless, this approach identified the abundance of numerous potential pathogens (Supp Fig 4), including *Campylobacter jejuni*, *Enterococcus faecium, Shigella sonnei, and Bordetella pertussis*. Additionally, *Helicobacter pylori* – a known risk factor for gastric cancer ^21^– was detectable. While culturing or qPCR-based validation is needed, the ability to detect cancer-associated microbes could potentially indicate another use of wastewater sequencing in tracking prevalence diseases in a population.

**Figure 4:**
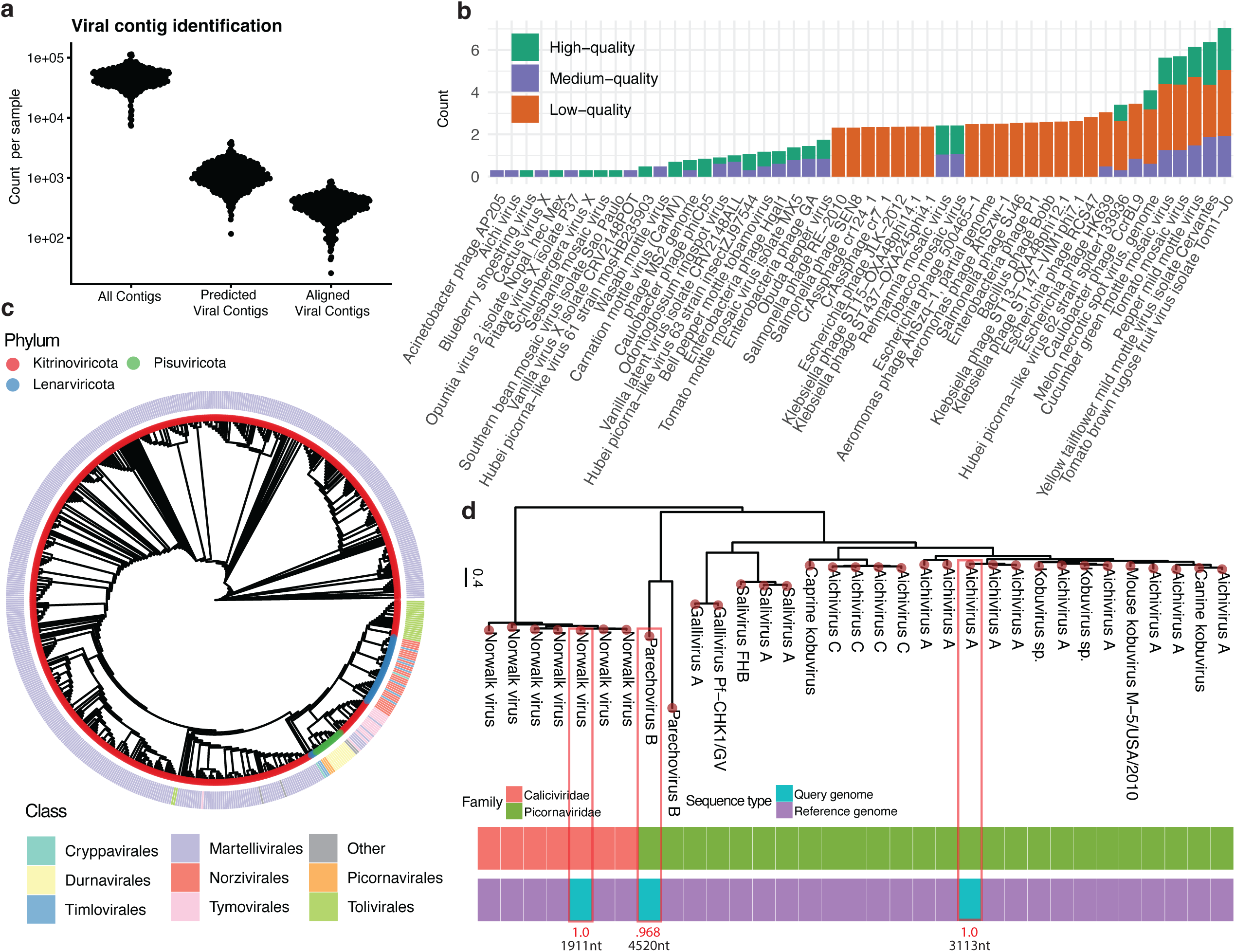
The assembled universe of viral content in wastewater. An alternate view of viral taxonomic composition within wastewater as generated by *de novo* assembly and comparison to reference databases. A) The total number of contigs assembled (per sample), the number annotated as low, medium, or high quality viruses, and the number aligning to a RefSeq viral genome at >90% identity. B) For low, medium, and high quality contigs, the top 25 most common annotations within each category (by RefSeq alignments). C) A cladogram (where branch lengths are normalized and not representative of evolutionary distance) of RNA viral life within this dataset. The tip points are labeled according to phylum, whereas the outer ring designates viral class. D) A phylogeny placing assembled enteric viral genomes against their genbank references. Red values are the FastTree2 Sh-like local support values for each query contig, black numbers underneath are said contigs’ lengths.

### The wastewater virome and its relevance to human health

The wastewater virome demonstrated less consistency across sites than bacteria, with most observed viruses being low prevalence (Supp Fig 5A-C). Only four high-prevalence viruses were identified. These prevalent viruses were associated with food (e.g., *Tomato brown rugose fruit virus, Tomato mosaic virus, Pepper mild mottle virus, Cucumber green mottle mosaic virus*), which may further indicate the ability to track dietary patterns via bulk sequencing (Supp Fig 5D). In low prevalence viruses (Supp Fig 5E) we also detected the expression of many phage genes; these were concordant with the bacterial clades identified (e.g., *Faecalibacterium, Enterobacteria*). Of particular relevance to human health was the identification of two *norovirus* strains and an *Aichivirus* (a potential cause of gastroenteritis) in a small subset of samples in the hospitals and the dormitories.

**Figure 5:**
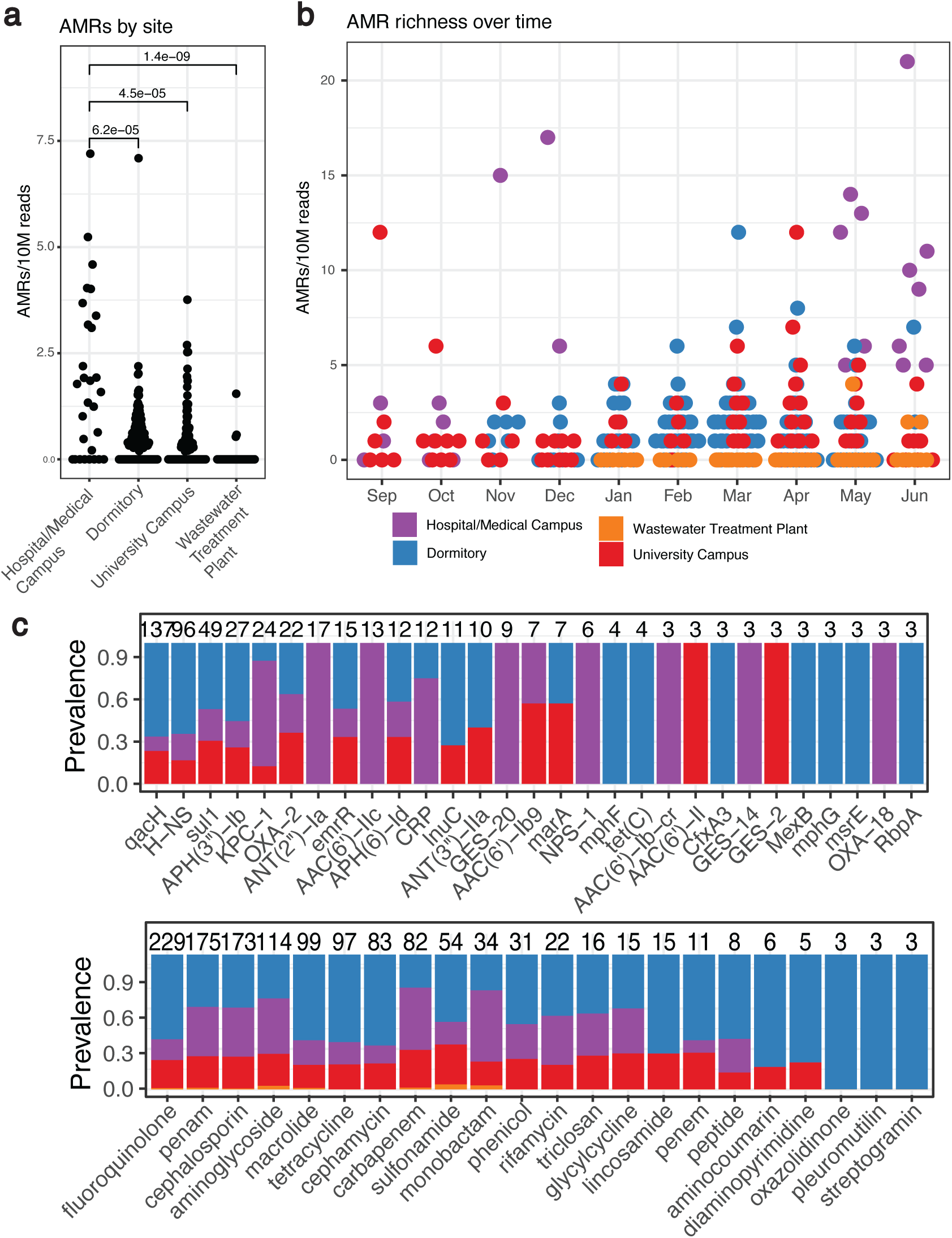
The Antimicrobial Resistance (AMR) landscape of wastewater across time and space. A) The total number of called AMR genes across different sample types per 10 million reads, with p-values deriving from Wilcoxon tests. B) The total number of AMR genes identified (per 10 million reads) across time. C) The top distribution of specific AMR genes and resistance mechanisms across all sample types.

Given the potential for spurious alignment in viral short read classification, we implemented a *de novo* assembly based approach for comparison. Assembly and viral contig classification provided another view into the diversity of wastewater and revealed an uncharacterized world of viral life. Of the 21.5 million contigs assembled, 527,144 passed our quality thresholds for being considered potentially viral (2.5% per sample, on average, Supp Table 4). As others have reported, the wastewater virome is poorly documented; on average, in a given sample, only 30% of low, medium, or high quality viral contigs aligned to complete viral genomes in RefSeq at >90% nucleotide identity (Fig 4A).

Taxonomic annotations for contigs that did align to RefSeq were generally concordant with those identified by short read classification (Fig 4B), though in some cases assembly provided increased taxonomic precision (Supp Table 4). For example, while short classification identified multiple animal-associated herpesviruses, only *Human herpesvirus 5* was identified by contig classification for ten contigs from discrete samples. Many of the most prevalent species by ANI were phages that ostensibly targeted human-relevant genera (e.g, *Enterobacter, Klebsiella, Shigella*), again indicating the potential of phage-expression-based biomarkers for bacterial pathogen prevalence. We additionally identified contigs aligning to *Norovirus* and *Aichivirus* reference strains. We also assembled one putative SARS-CoV-2 genome fragment with 99.6% identity to the Wuhan-Hu-1 reference, however it was extremely short (250 bases), and therefore potentially spurious.

We further explored the phylogeny of the wastewater RNA virome. We filtered for long viral contigs (>1000bp) and identified single-copy genetic features (found in at least 50 viral genomes each) shared across them (see *Methods*) to construct a cladogram to cluster organisms with similar genomes (Fig 4C). We grouped these long contigs with reference genomes based on genetic domain overlap to generate taxonomic annotations to the family level. We additionally provide a phylogenetic tree (i.e., with non-normalized branch lengths, Supp Fig 6). We found three different RNA viral phyla which clustered together on the resulting tree (*Kitrinoviricota, Lenarviricota, Pisuviricota*), though the tree building process filtered out many contigs corresponding to other phyla yet lacking highly prevalent genetic domains. These phyla comprise a cross-section of RNA viruses that infect hosts across the tree of life, from fungi (e.g., the class *Cryppavirales*) to mammals (e.g., *Picornavirales*) to plants and insects (e.g., *Tolivirales*) to bacteria (e.g., *Norzivirales*). Notably, this indicates the presence of fungi in wastewater, which we do not investigate in this study but also are highly relevant to host health.

From here, we aimed to further confirm the putative enteric viral genomes we had identified by both RefSeq alignment and domain-based taxonomic classification. We placed the members of families *Caliciviridae* and *Picornaviridae* on a phylogenetic tree (Fig 4D) with their most closely related GenBank references. The three queries – a low-quality *Norwalk virus* (i.e., *Norovirus*) contig, a low quality *Parechovirus B* contig, and a medium quality *Aichivirus A* contig – all were placed in the same clades as their appropriate reference. This indicates that even poor quality assembled viral genomes can be used for monitoring pathogens in wastewater with bulk RNA sequencing.

### The antimicrobial resistance and functional landscape of wastewater

We next characterized the functional landscape of wastewater, focusing on the expression of antimicrobial resistance (AMR) genes. The expression of AMR genes additionally varied across sites and time (Supp Fig 7, Supp Table 5). Hospital/medical campus outflow contained the greatest number of predicted AMR genes on average, more so than any locale (Wilcoxon p-value < 0.05 in all comparisons, Fig 5A), with wastewater treatment plants containing the least. AMR genes also displayed potential seasonality, with increased numbers identified in the early spring months (Fig 5B). We hypothesize that this could be due to the increased student population on campus or, potentially, an increase in antibiotic prescriptions. The top AMR genes found were qacH, H-NS, sul1, APH(3”)-lb, and KPC-1 (Fig 5C). The hospital samples had high numbers of KPC-1, AAC(6’), ANT(2”)-Ia, GES-20, NPS-1, and CRP. The top antibiotic classes with resistance overall were fluoroquinolones, penams, cephalosporins, aminoglycosides, and macrolides.

## Discussion

In this longitudinal wastewater microbial census, we integrate targeted and untargeted approaches to detail the bacterial, viral, and functional landscape of samples from multiple areas within Miami-Dade County across two years. By integrating multiple, redundant bioinformatic approaches, we show the site-specific and shared features that indicate the potential for wastewater monitoring to be used for 1) pathogen surveillance, 2) community epidemiology, including diet and gut health, 3) pollutants, such as metals and fertilizers, and 4) tracking geospatially varying antimicrobial resistance, among other functionally-relevant signatures. With the increased integration of wastewater sequencing and geospatial epidemiologic data, we can identify increasingly precise signatures for these and other features, potentially replacing onerous and costly public health surveillance methods in the long run.

Targeted and untargeted analyses are complementary and can provide strategic direction for public health policy. Targeted ARTIC sequencing, for example, can identify extremely low abundance (parts per billion) viruses that require many rounds of amplification via PCR to be able to sequence. However, it is limited in its ability to only capture a single organism per set of primers. Bulk sequencing, on the other hand, can theoretically capture information from all domains of life living in wastewater. Given that wastewater is an aggregate of environmental and human biology from a single geography, it stands to reason that microbial signals in wastewater could be used as a one-stop-shop for public health surveillance. From heavy metal levels (e.g., lead, copper, fertilizer runoff) to gut health to cancer clusters, theoretically integrated microbial surveillance could capture it all in one place for low cost, with limited privacy concerns. In this study, for example, we both tracked SARS-CoV-2 VoC levels over multiple years and simultaneously were able to identify abundance changes in human commensal bacteria, pathogens, and enteric viruses. We additionally highlighted the ability of wastewater sequencing to capture human-relevant microbial functional elements, like AMR genes.

An added benefit of the wastewater surveillance documented here, specifically, is the continuous monitoring of geographically distinct sites serving different community sizes. We observed variation in microbial composition and consistency across represented population size, for example, observing low variability in wastewater treatment plant microbiomes compared to dormitories. It stands to reason that temporal variation in major environmental pollutants might be best monitored at treatment plants, whereas isolated outbreaks (e.g., gastroenteritis) might be better tracked at locations housing smaller populations.

Of course, our approach is not without drawbacks. Taxonomic classifiers often do not perform well on environmental data, and a large percentage of our reads went unclassified; this will be improved as reference genome databases continue to expand. Additionally, annotation of viruses is extremely challenging. Applying multiple methods (e.g., multiple databases and algorithmic approaches, and/or long reads) and seeking concordance between them. Algorithmic improvements are also needed in order to increase confidence in decisions made based on wastewater surveillance.

Additionally, this study is primarily focused on characterizing geospatially-determined ecology; we do not have data on, for example, heavy metals or other large-scale public health hazards. Future research needs to combine extensive wastewater datasets with existing observational and epidemiological cohorts (e.g., the National Health and Nutrition Examination Survey, NHANES) across diverse zip codes. Only through such comprehensive studies can the true potential and value of wastewater surveillance be effectively demonstrated and quantified.

Overall, this dataset demonstrates the breadth of potential for the pragmatic utilization of wastewater as a tool for public health interventions. By characterizing the ecology and its variation across time and space, we aim to lay the groundwork for building a dataset of taxa of interest to be monitored with bulk RNA-sequencing alongside targeted approaches for pathogens of import. That said, wastewater monitoring requires microbial variation explained by human-relevant events (e.g., pandemics, nutritional status) to be divergent from variation explained by natural fluctuations in community ecology; further effort will have to be exerted to hone in on the most practical biomarkers, as well as the methods for detecting them. Downstream, though, we propose that a national, or even global, semi-automated surveillance system would serve as a first line of defense against future pandemics, contaminant buildup, and other public health crises.

## Methods

### Study design and IRB approval

The goal of the study was to evaluate associations between clinical data and wastewater SARS-CoV-2 RNA levels in matched populations. Starting the Fall 2020 semester, to optimize on-campus safety against the spread of COVID-19, the University of Miami embarked on an aggressive clinical tracking program based upon testing, tracking, and tracing (3-T). For students who lived or commuted to campus, the University required weekly or bi-weekly testing. Employees were tested on a random basis. Documented clinical data were thus available on a building basis at the University of Miami dormitories, for the University campus for both student and employee populations who resided or commuted to campus, and at the University of Miami hospital. In addition, a subset of positive clinical samples (both students and employees) were amplified for SARS-CoV-2 variants and sequenced at a University of Miami laboratory (Oncogenomics Shared Resource, OGSR). The OGSR ran both the clinical and wastewater samples reported in this study providing for consistency in the detection between both data sets. The sharing of clinical data for internal research purposes was approved by the University of Miami IRB (MOD00003977). In addition, at the county level, the Florida Department of Health documented in a publically available database positive COVID-19 detections by zip code. Given the availability of clinically-based data, the wastewater sample collection program was designed to provide a population match between clinical data and populations contributing wastewater to a specific wastewater sampling location.

### Sample collection

Samples were collected weekly from September 30, 2020 through September 21, 2022. Sampling sites included manholes from residential dormitories (14 sites), sites representing major portions of campus (4 sites, 3 corresponding to the main residential campus and 1 corresponding to the university marine campus with no residences), from laboratory/administrative buildings at the medical campus (2 sites), from the University hospital (2 sites), and from a regional wastewater treatment plant (Central District) serving a population of 830,000 from Miami-Dade County. In addition, at three sites both grab and composite samples were collected accounting for the balance of the 26 sampling sites. Between September 2021 and January 2022, samples were collected two-times per week at the dormitory sites to provide additional data for on-campus mitigation measures.

Composite samples were collected at the building and cluster scale using either an ISCO 6712 autosampler (time-paced) or at the community scale wastewater treatment plant using an HACH autosampler (flow-paced). All other samples were collected using a grab sampling technique, due to limited availability of multiple autosamplers. Sample collection volumes were 2 liters with the first liter being used for water quality analyses in the field (pH, water temperature, dissolved oxygen, specific conductivity, and turbidity). Additional details about sample collection and basic water quality data have been published earlier (reference Sharkey et al. 2021, Zhan et al. 2022, Babler et al. 2022, Babler et al. 2023, and Solo-Gabriele et al. 2023. The DOIs are given above.)

### Primary concentration and downstream processing of wastewater

Upon collection, wastewater was transported to the Biospecimen Shared Resource Laboratory at the University of Miami on ice and immediately succumbed to pretreatment for primary concentration of suspended solids. Wastewater samples had [10>6 genomic copies(gc)/L] human coronavirus-OC43 added as a recovery tool for quantitative PCR assessment, occurring prior to targeted sequencing. Additionally, 51% w/v magnesium chloride was added as well as drops of 10% Hydrochloric Acid (pH reduction down to 3.5-4.5) to assist with altering the chemical charge of ambient viral particles found in wastewater to positive. The primary concentration method employed was Electronegative Filtration (ENF) (Sharkey et al. 2021, Babler et al. 2022, Zhan et al. 2022, Babler et al. 2023, Solo-Gabriele et al. 2023) using HAWP mixed cellulose ester membranes (Millipore Sigma #HAWP04700, 47 mm diameter, 0.45 µm pore size) which are negatively charged, thus requiring the necessity for the pretreatment to aid in binding affinity between the viral particles and membranes. Via the use of vacuum filtration, electronegative HAWP membranes captured wastewater suspended solids, and variable sample volumes (20-150 mL, depending upon turbidity and water quality parameters) were filtered until clogging/filter saturation to create concentrates. HAWP filter membranes containing a saturated layer of suspended wastewater solids were folded in on themselves four times, and placed immediately within 1.5 mL 1X DNA/RNA shield within 5 mL microcentrifuge tubes. Three filter concentrate replicates were generated per wastewater sample collected over the course of the study period; each replicate was sent to a separate laboratory, the Center for AIDS Research (CFAR) Laboratory (on ice), the OGSR Laboratory (on ice) both at the University of Miami, and at Weill Cornell Medicine (WCM) (on dry ice).

Three workflows were implemented to extract SARS-CoV-2 from wastewater which occurred in separate laboratories to standardize and achieve overlapping analyses following extraction. These processes included quantitative PCR, ARTIC analysis, and bulk RNA-sequencing. At the CFAR laboratory the Zymo Research QuickRNA-Viral Kit, with a modified protocol using 250 µL wastewater concentrate - discussed in detail within Babler et al. 2023 to reduce inhibition - was utilized as input for Volcano 2nd Generation-qPCR (V2G-qPCR). This in-house created assay is equivalent to that of RT-qPCR (Babler et al. 2022) but bypasses the need for the initial reverse transcription step as the DNA polymerase utilized within the reaction can read both RNA and DNA templates, thus reducing the total reaction time and allowing for additional analyses of other targets within the same timeframe (i.e., normalization parameters, control parameters) (Sharkey et al. 2021, Babler et al. 2022, Babler et al. 2023). To assess for inhibition within RNA samples, 30 µL of HIV RNA was added to each 10 µL eluate of wastewater RNA, and assessed alongside a weekly generated water control (10 µL Nuclease-Free water + 30 µL HIV RNA) by V2G-qPCR; following amplification, the Cq values were compared and if the wastewater sample Cq values measured within ±2 cycles of the water control, samples were considered uninhibited (Babler et al. 2023).

The choice for determining which samples were ultimately brought through ARTIC analyses, via collaborative communication between CFAR and OGSR, were based on the gc/L measurements determined by V2G-qPCR taking into consideration the overall recovery and inhibition as determined by the controls. This was because routine extraction and multiple V2G-qPCR assessments occurred within the CFAR laboratory weekly, wherein the filter replicate sent to the OGSR following concentration was kept in −80 C until extracted in batches for specific analyses.

### Targeted sequencing of SARS-CoV-2

Libraries were synthesized using the NEBNext® ARTIC SARS-CoV-2 FS Library Prep Kit (7658L) following the Express Protocol and employing the V3 primer set throughout. Eight microliters of MagMAX TNA was used as input without quantification nor assessment of RNA integrity. Finished libraries were pooled volumetrically with final pools of 96 libraries cleaned using 0.9x AMPure bead cleanup (Beckman A63882). Pools were sequenced on a Illumina NextSeq 500 using Mid-Output 150 cycle flow cells (130M clusters, 20024904) run as 76:8:8:76 cycles with 3% PhIX (FC-110-3001). Generic Bcl2fastq was run in Illumina BaseSpace.

### Detection of SARS-CoV-2 variants in ARTIC data

Broadly speaking, the pipeline for variation detection involved (1) identification of reads aligning to the SARS-CoV-2 genome with Kraken2, (2) aligning to the Wuhan reference and filtering samples based on breadth and depth of alignment coverage, (3) trimming primers, (4) calling variants using an ensemble approach, (5) annotating mutations, and (8) estimating VOC lineage abundances. All software was run with the default settings unless otherwise specified.

We used Kraken2^22^ (V2.1.2) running the default settings to taxonomically classify short read sequencing data for the ARTIC samples. We used a custom Kraken2 database that included the SARS-CoV-2 Wuhan reference (GCF_009858895) as well as an assortment of viral, archaeal, bacterial, fungal, protozoan, and mammalian genomes (available upon request). The reads assigned exclusively as SARS-CoV-2 are then filtered into individual files using seqtk (V1.3-r106). Alignment to the Wuhan reference is done with bwa (V0.7.17-r1188), and the alignment is then sorted and indexed with sambamba. We then trim primers with ivar (V1.13) and generate coverage statistics afterwards with the bedtools genomecov (V2.30.0) command and mosdepth (V0.3.3). We filter out samples that did not have at minimum 10x mean coverage per amplicon across at least 73 of the 98 amplicons (roughly 75%) that were targeted by the ARTIC protocol.

For variant calling, we used an ensemble approach, combining the output of lofreq (V2.1.5), and iVar, only the union of calls found in both approaches and computing the mean variant allele frequency between the two. We used VEP (V104.3) to annotate mutations and estimate VOC lineage abundance with Freyja (V1.4.2), using the latest available VOC database, provided by the software.

### Bulk RNA-sequencing

After concentration via ENF, the filter concentrates were preserved in 1.5 mL of 1X DNA/RNA Shield (Zymo Research), transported on dry ice, and remained stored at −80°C until further processing at Weill Cornell Medical College. Nucleic acid extraction took place using the DreamPrep NAP workstation (TECAN), a liquid-handling automation device, and the Quick-DNA/RNA Viral Magbead Kit (Zymo Research) using 400 µL of wastewater concentrate, following manufacturer’s recommendations.

HudsonAlpha Discovery (Huntsville, AL) performed all sequencing library preparation on the generated nucleic acids. The nucleic acid samples underwent purification using the RNA Clean and Concentrator Magbead Kit with DNase digestion (Zymo Research), following the manufacturer’s recommended protocol on the Biomek i5 automated workstation (Beckman Coulter). This process resulted in an 18 µL elution of RNA in nuclease-free water. The LabTouch GX Touch (Perkin Elmer) nucleic acid analysis system quantified the RNA, with samples prepared according to the manufacturer instructions.

After normalizing RNA concentrations, sequencing libraries were generated using the NEBNext rRNA Depletion Kit and (Human/Mouse/Rat) NEBNext Ultra II DNA Library Prep Kit (New England Biolabs) for Illumina. The KAPA Library Quantification Kit (Roche) was employed to quantify the resulting libraries, all samples were brought to 20 cycles of PCR at the final step. All libraries were next evaluated and pooled at equimolar concentrations. Sequencing was carried out on the NovaSeq 6000 platform (Illumina) with an S4 flow cell, using a read length of 2 x 151 bp in paired-end configuration.

### Metatranscriptomic quality control and short read taxonomic profiling

We submitted all short read metatranscriptomic sequencing data first to a quality control pipeline, predominantly using the bbtools suite (V38.92)^23^. Clumpify (parameters: optical=f, dupesubs=2,dedupe=t) was used to group overlapping reads, and bbduk (parameters: qout=33 trd=t hdist=1 k=27 ktrim=“r” mink=8 overwrite=true trimq=10 qtrim=’rl’ threads=10 minlength=51 maxns=-1 minbasefrequency=0.05 ecco=f) was used to deduplicate reads and remove adapters, accounting for duplicates generated by the PCR-amplication process. Potential human contaminating reads were removed by bowtie2 (V2.4.4, parameters: --very-sensitive-local) alignment to the HG38 human genome assembly^24^. Any samples remaining with uneven numbers of reads were repaired with bbtools’ repair function. Finally, tadpole (parameters: mode=correct, ecc=t, ecco=t) was used to correct sequencing errors.

Quality-controlled reads were run through different taxonomic classification approaches: 1) Kraken2 (V2.1.2) against all of the complete genomes for microorganisms on RefSeq, 2) xtree (V0.92i) against the Genome Taxonomy Database (GTDB) r207, xtree against the dataset of complete GenBank viral references, 4) and MetaPhlan4 (V4.0.4) against its default database. Xtree is a k-mer based aligner that generates coverage statistics (global, which corresponds to total genome coverage, and unique, which corresponds to reads mapping to a specific genome in the database). We filtered for bacterial genomes with 1% global and 0.5% unique coverage. We filtered for viral genomes with 20% global and 10% unique coverage. Relative abundances for xtree were computed by dividing total aligned reads to a given genome over the total number of reads aligning to all genomes. MetaPhlAn4 computed relative abundance automatically. We used bracken to compute relative abundances for the kraken2 output. All the figures in the text were generated with the xtree relative abundances, unless otherwise specified.

The bacterial phylogeny generated in Figure 2A was generated by subsetting the GTDB taxonomic tree to the species identified in the metatranscriptomic data. Family-level representatives were selected at random. Their abundance, reported in the circular heatmap, was computed by averaging the relative abundance of all species in that given family.

### Microbial Association Study

The Microbial Association Study (MAS) reported in Fig 3F and Supp Fig 3 was executed via a linear modeling approach. Relative abundances were log-transformed with the smallest non-zero abundance value being added to the entire matrix beforehand. The following model was fit for each bacterial species:

> *log(microbial_feature_abundance_n_) ~ environmental_feature_i_*

The dependent variable is the log transformed abundance of any given bacteria and the dependent variables are the features described in Supp Fig 3. P-values were adjusted via the Benjamini-Hochberg procedure, and a q-value cutoff of 0.05 was used to gauge significance.

### De novo assembly, viral classification, and AMR/Open-Reading-Frame identification

Quality-controlled reads were *de novo* assembled using MetaViralSPAdes. Open-reading-frames were identified and annotated with Bakta. Antimicrobial resistance genes and virulence factors were identified with Abricate (V1.0.1). We can CheckV (V0.8.1) on all assembled contigs. Those that were annotated as low, medium, or high quality were selected for further analysis and considered potentially viral. All tools were run with the default parameters.

We took two approaches for taxonomically characterizing viral contigs: alignment-based HMM-based. For the former, we aligned all contigs back to the RefSeq viral database using BLAST V2.12.0+ ^25^. We modeled the alignment parameters (-max_target_seq 25000, -perc_identity 90 - outfmrt “6 std qlen slen qcovs”) based on published methods^26^; the scripts from this manuscript are available at https://github.com/snayfach/MGV/tree/master/ani_cluster. We report in Fig 4B the top hit for each query contig, stratified by its CheckV-determined quality.

The second approach involved searching viral contigs against a database of single-copy, genetic-features, in the form of Hidden Markov Models (HMMs). This approach is also modeled on existing methods^27^. To generate our contig database, we used hmmer (V3.3.2) to search all complete viral contigs on GenBank against the Pfam and TIGRFAM HMM databases, selecting for only single copy annotations in the output^28, 29^. We searched assembled viral contigs that were over 1000 nucleotides in length against these markers (e-value cutoff of 0.1), again filtering out any that came up as multi-copy. We assigned putative taxonomic classifications based on the closest hit in terms of HMM overlap with GenBank viruses. We used the predicted phylum to filter out viruses that were non-RNA.

For each predicted RNA virus, we further searched for a minimum set of single HMMs within those annotated that were shared by at least 50% of the contigs. The sequences of these HMMs were concatenated for each contig and multiple sequence alignments were carried out using FAMSA (V2.1.2)^30^. The resultant alignments were placed in a phylogeny with GenBank reference genomes (that shared overlap in the same HMMs) using FastTree (V2.1.11, parameters: -fastest

-pseudo). The cladogram (with normalized branch lengths, so showing relatedness and not evolutionary distance) in Fig 4C is this tree with the GenBank references filtered out. The phylogeny in Fig 4D is this tree, filtered to a subset of contigs, with the references kept in.

### Additional software

All analysis was done in RV4.1.3^31^. Additional packages used include: ggtree V3.7.2^32^, ggplot2 V3.4.2^33^, the tidyverse V2.0.0^34^, ComplexHeatmap V2.10.0^35^, UpSetR V1.4.0^36^, ComplexUpSet V1.3.3^37^, phytools V1.5-1^38^, ape V5.7-1^39^, umap V0.2.10.0^40^, ggbeeswarm V0.7.1, tidytext V0.4.1^41^, circlize V0.4.15^42^, vegan V2.6-4^43^, broom V1.0.4^44^, reshape2 V1.4.4^45^, and ggpubr V0.6.0^46^.

## Data availability

In compliance with the NIH RADx-rad Data Coordination Center (DCC) requirements, the raw sequencing data was submitted to the Sequence Read Archive (SRA). The wastewater samples were annotated with the rich metadata and the sequencing information (fastq files) was included in the submission. The submitted data can be found in the SRA under the accession PRJNA946141.

Furthermore, the metadata associated with the wastewater samples’ sequencing data was extracted from the Illumina operational files, validated, organized, and submitted to the NIH data hub via DCC [https://radx-hub.nih.gov/home], where the SF-RAD data is associated with the dbGaP study accession phs002525.v1.p1.

The metadata standards specifications used to describe the data were developed in collaboration with the SF-RAD members and the DCC and formally defined and registered at FAIRsharing.org.

## Code availability

The software for the viral HMM-based taxonomic classification and phylogeny construction is available at https://github.com/b-tierney/vironomy. All other software used were published tools (which we report the parameters and implementations for in the other Methods sections). Scripts used for plotting and slurm-based job management are available at https://github.com/b-tierney/radx-wastewater-scripts/tree/main.

## Data Availability

In compliance with the NIH RADx-rad Data Coordination Center (DCC) requirements, the raw sequencing data was submitted to the Sequence Read Archive (SRA). The wastewater samples were annotated with the rich metadata and the sequencing information (fastq files) was included in the submission. The submitted data can be found in the SRA under the accession PRJNA946141. Furthermore, the metadata associated with the wastewater samples' sequencing data was extracted from the Illumina operational files, validated, organized, and submitted to the NIH data hub via DCC [https://radx-hub.nih.gov/home], where the SF-RAD data is associated with the dbGaP study accession phs002525.v1.p1. The metadata standards specifications used to describe the data were developed in collaboration with the SF-RAD members and the DCC and formally defined and registered at FAIRsharing.org.

## Acknowledgments

This study was financially supported by the National Institute on Drug Abuse of the National Institutes of Health (NIH) under Award Number U01DA053941. The content is solely the responsibility of the authors and does not necessarily represent the official views of the NIH.

## Competing interests

BTT is compensated for consulting with Seed Health and Enzymetrics Biosciences on microbiome study design and holds an ownership stake in the former. CEM is a co-Founder of Onegevity and Biotia. No entity listed here was involved in funding or advising the contents of this study.

## Supplemental Figures and Tables

**Supplemental Figure 1:**
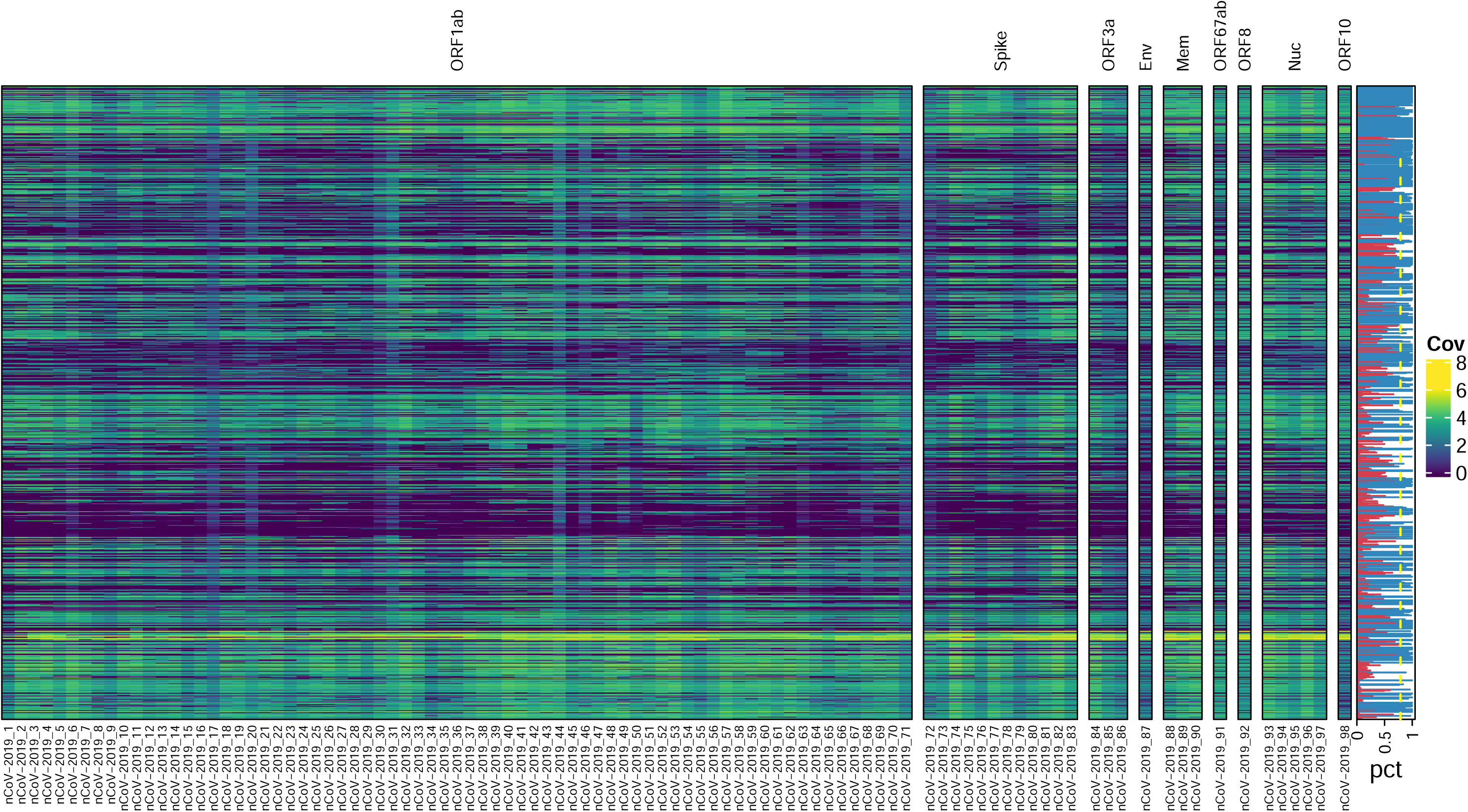
Coverage of SARS-CoV-2 Wuhan reference genome in all wastewater samples.

**Supplemental Figure 2:**
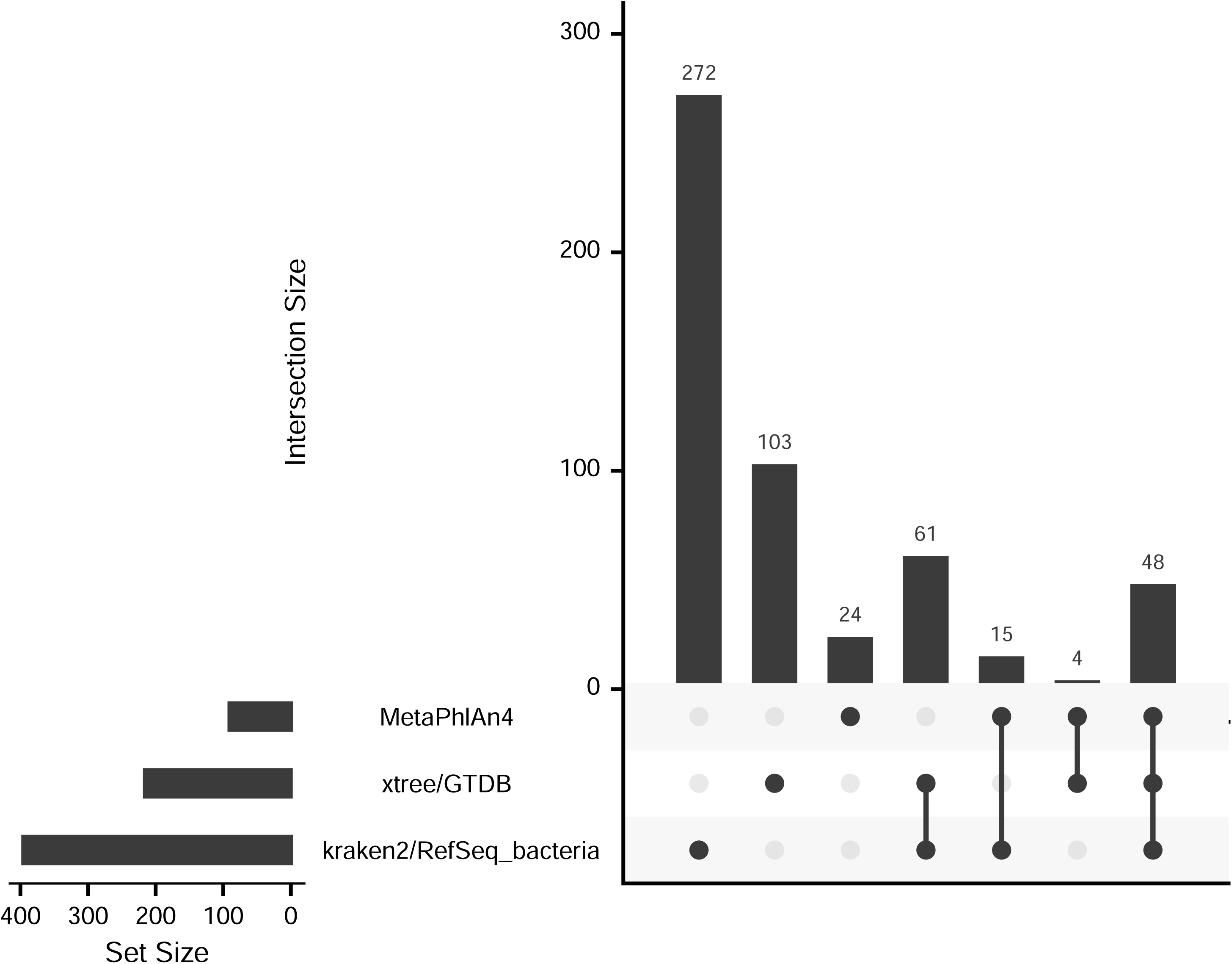
Overlap between identified bacterial families in metatranscriptomic data by different classification approaches.

**Supplemental Figure 3:**
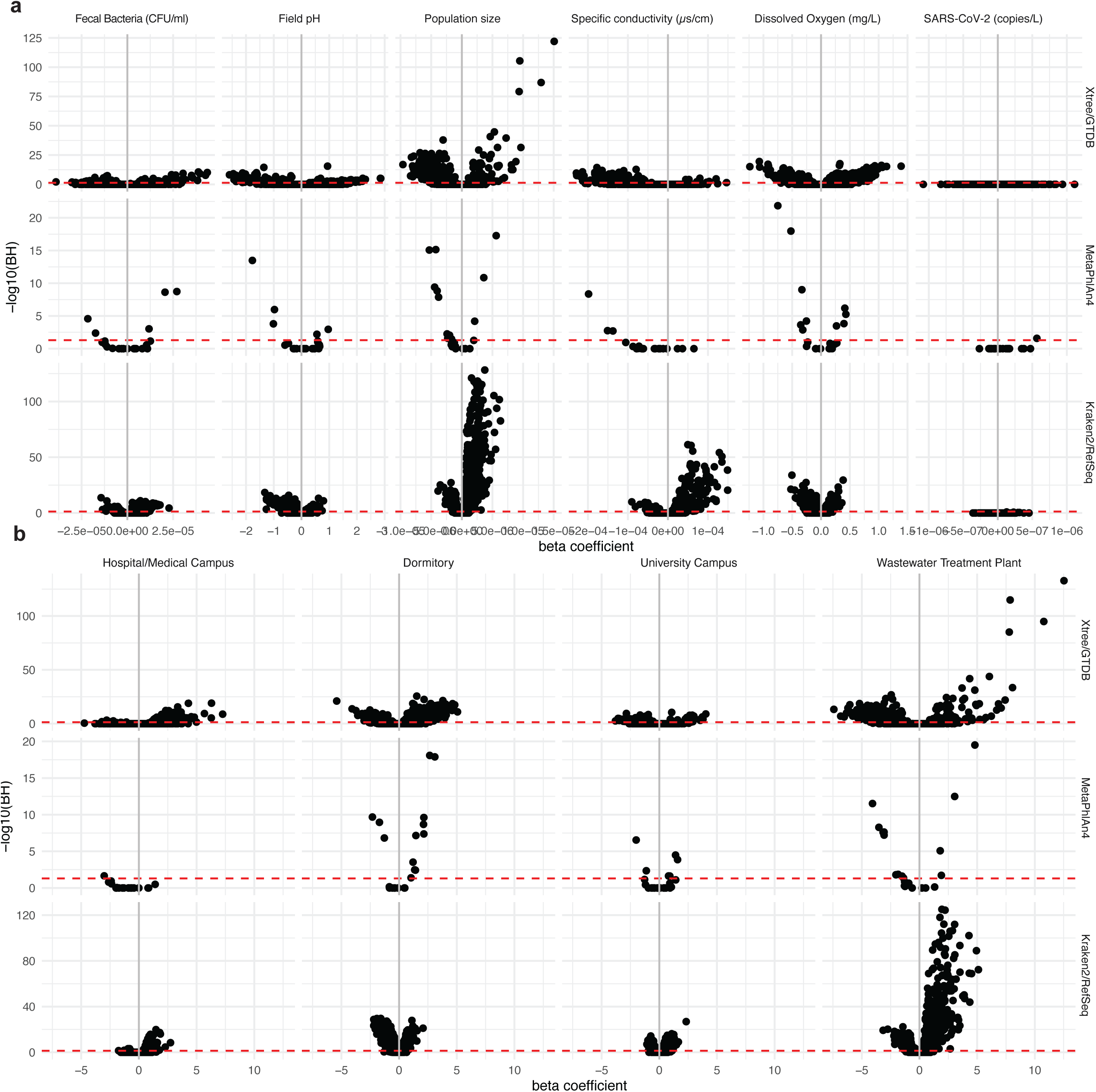
A Microbiome Association Study between bacterial expression and environmental features. Volcano plots depicting the number of associated features and direction therein for different variables. X-axis is the beta-coefficient from the variable listed at the top of the plot. Y-axis is negative log(10) BH-adjusted p-value (i.e., q-value).

**Supplemental Figure 4:**
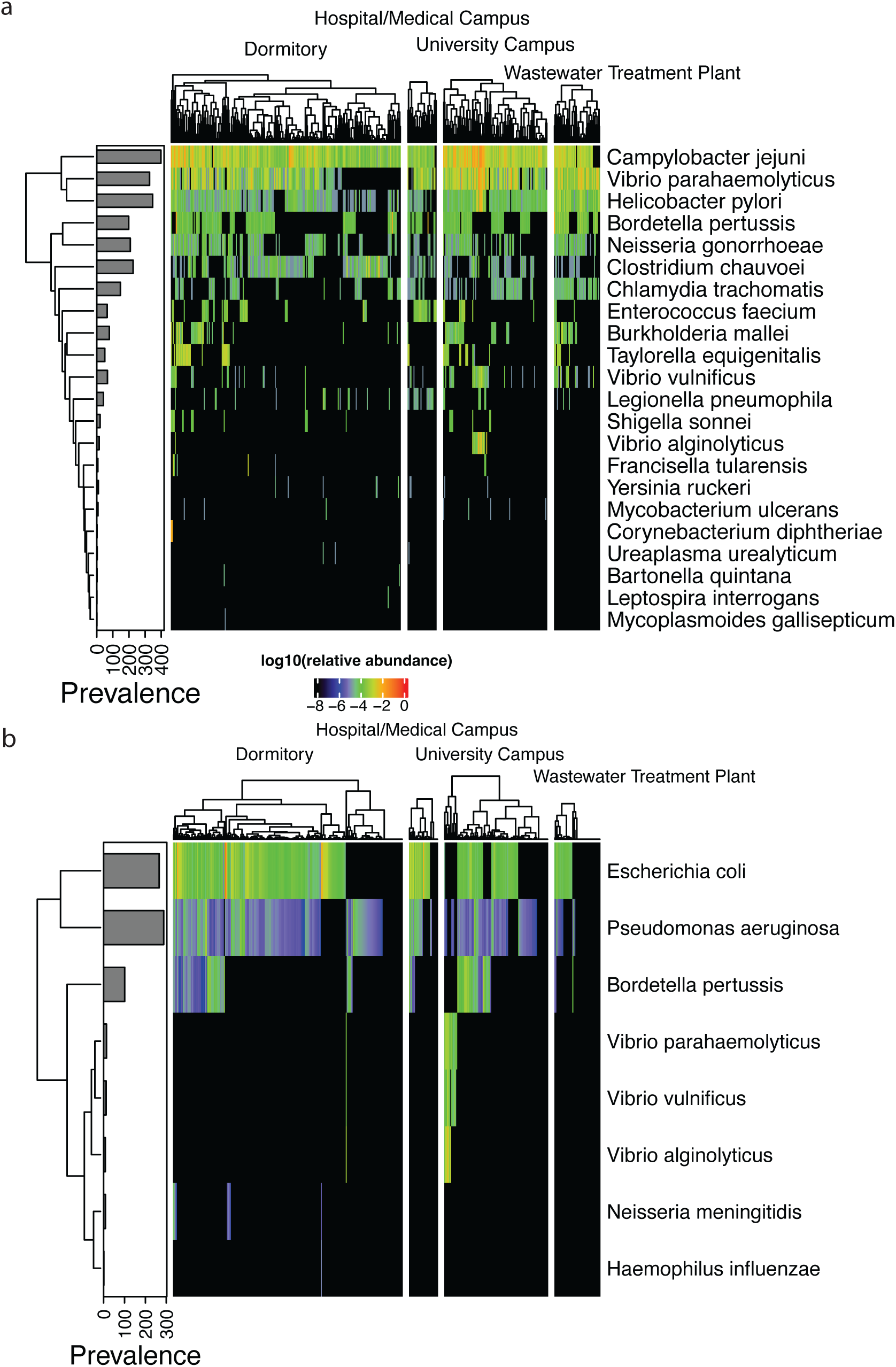
Putative, low abundance pathogens and cancer-associated bacteria as identified by A) Kraken2 and B) XTree.

**Supplemental Figure 5:**
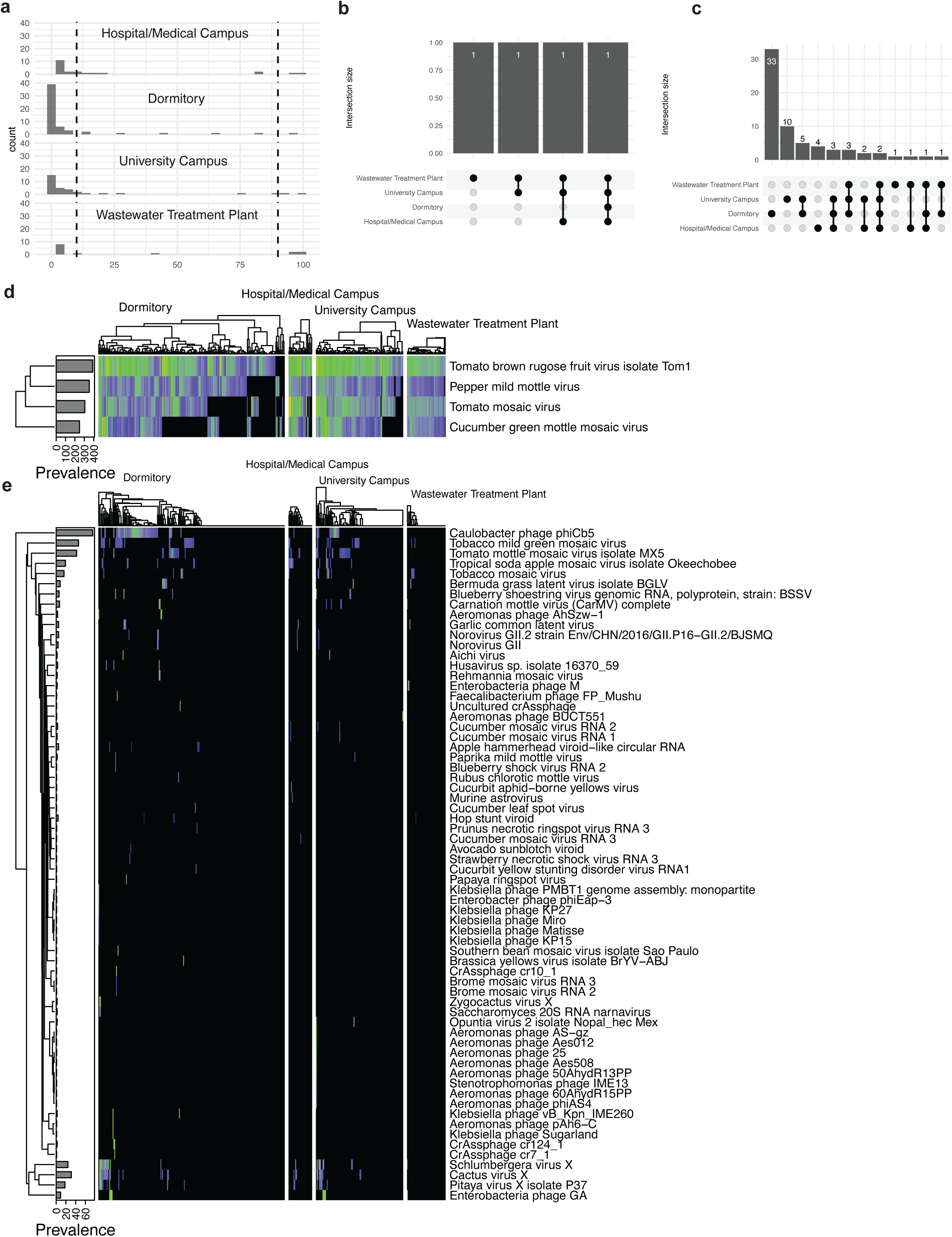
Viral identification by short read alignment. A) The prevalence of all identified viral species across different types of sampling locations. B-C) The overlap between high prevalence (B) and low prevalence (C) organisms across different sampling locations. D-E) The log10(relative abundance) of high-prevalence (D) and low-prevalence (E) viral species across all sites. These two panels display, at most, the union of the top 25 most abundant organisms across each sampling location and category (high or low prevalence).

**Supplemental Figure 6:**
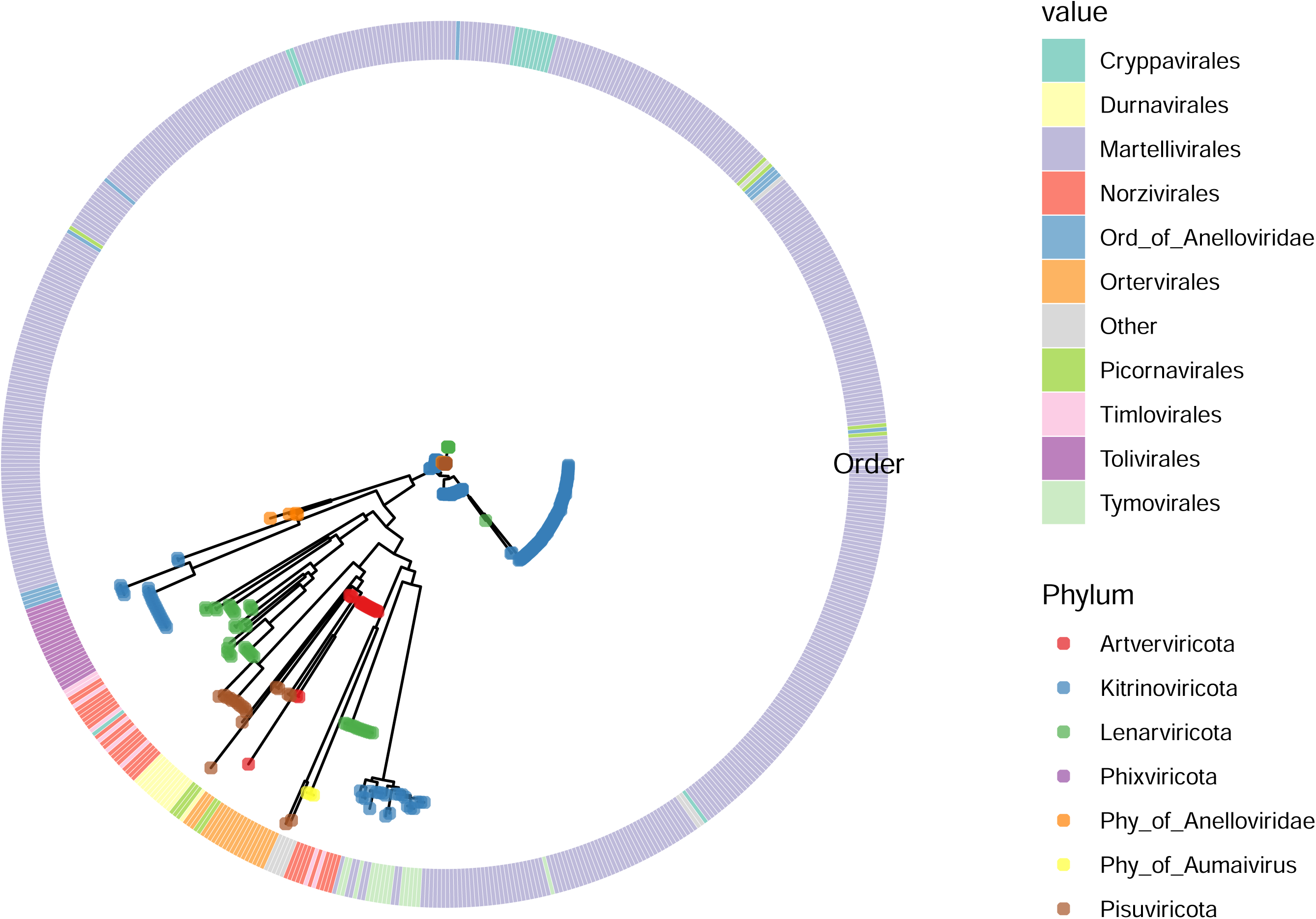
The phylogeny of assembled RNA viruses in wastewater.

**Supplemental Figure 7:**
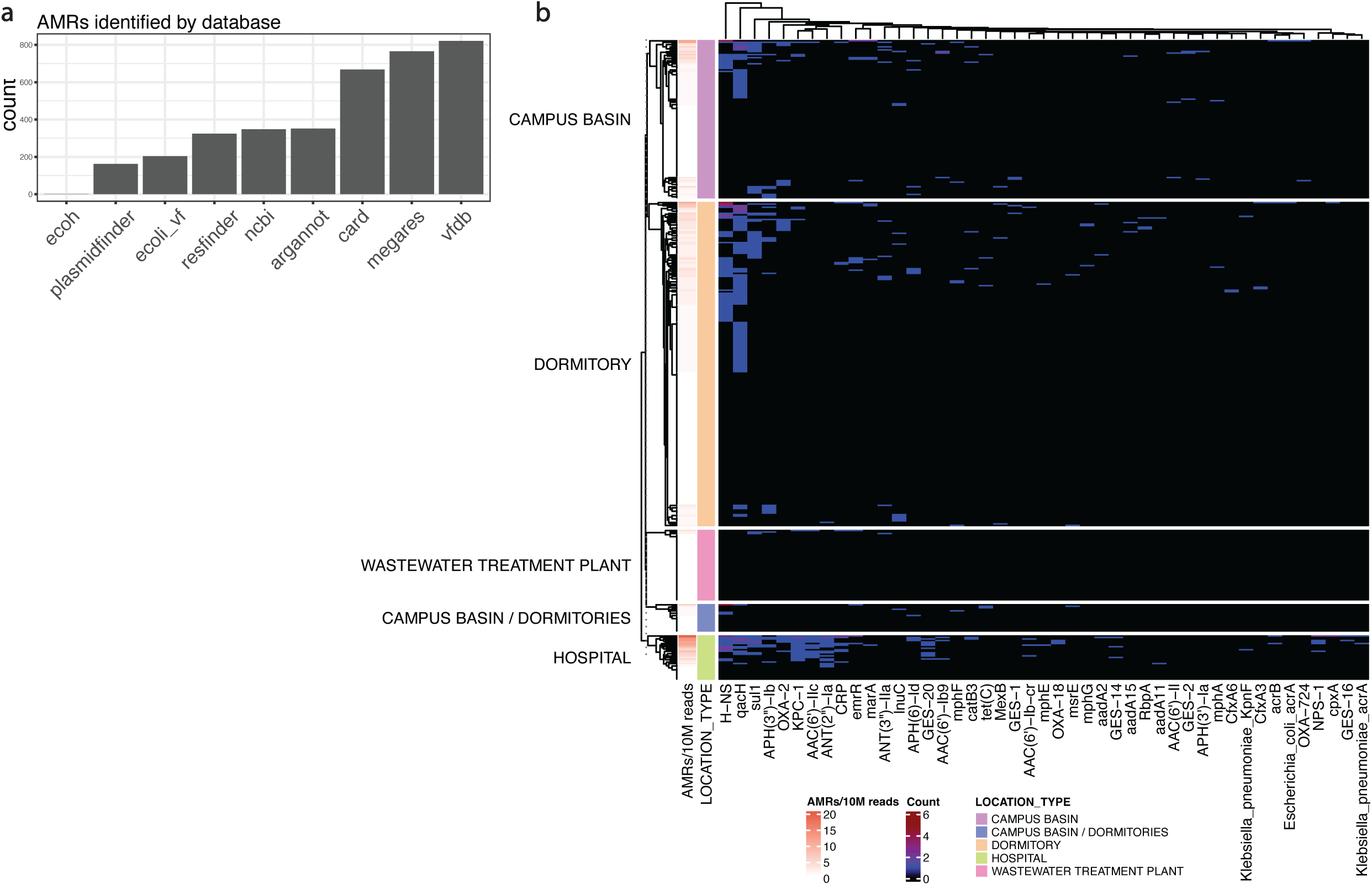
The antimicrobial and virulence landscape of wastewater. A) Antimicrobial resistance genes, plasmids, virulence factors, and other host-relevant genes as annotated by database. B) The expression of antimicrobial resistance genes across all sites and samples as annotated by CARD.

**Supplemental Table 1**: Results from SARS-CoV-2 VoC identification.

**Supplemental Table 2**: Complete set of taxonomic tables for all short read taxonomic classification approaches used in this manuscript.

**Supplemental Table 3**: Associations between microbial taxonomic features and wastewater characteristics.

**Supplemental Table 4**: Viral assembly analysis. Includes checkV quality scores and alignment-based classification of putative viral contigs as well as best-hit taxonomic annotations for RNA viruses used in Fig 4D.

**Supplemental Table 5**: Antimicrobial resistance and virulence gene data across all samples.

## Notes

### Author Declarations

The IRB of Weill Cornell Medicine gave ethical approval for this work.

